# A seven-day cycle in COVID-19 infection, hospitalization, and mortality rates: Do weekend social interactions kill susceptible people?

**DOI:** 10.1101/2020.05.03.20089508

**Authors:** Itay Ricon-Becker, Ricardo Tarrasch, Pablo Blinder, Shamgar Ben-Eliyahu

## Abstract

Seven-day cycles in numbers of COVID-19 new-cases and deaths are markedly evident in most public databases (e.g. Worldometer, ECDC), but it is unclear whether they reflect systematic artifacts of delays in information reporting/gathering, or have a more profound basis. To address this question we located 11 databases of US states that provide date- authenticated information (actual date of symptom onset and/or specimen collection, or actual hospitalization or death date) that reported more than 1,000 deaths each. Numbers of new cases showed a weekly cyclic pattern in 10 out of 11 states, commonly peaking on weekdays, 2-6 days after the weekend, corresponding with a reported median 5-day lag between infection and the manifestation of clinical symptoms. We postulate that this pattern emerges from interactions with different and/or extended social-circles during weekends, including increased inter-generational meetings, which in turn facilitate transfer of COVID-19 from younger people to older vulnerable individuals. Furthermore, we found weekly periodicity in hospitalizations in 2 out of 2 authenticated databases providing this information. Actual death date, which is more difficult to attribute to individual choice, and is expected to occur approximately 2-3 weeks following hospitalization, showed significant 7-day periodicity in 1 out of 11 states, and a trend in 2 additional states. If weekly peaks in new cases can be truncated by physical/social distancing, especially during weekends, the mortality of COVID-19 may be reduced, or at least hospitalization and mortality curves may be flattened.

**Significance Statement:** We believe our findings are significant, as it appears that it is difficult for people to grasp an ambiguous “price” for visiting their friends and family, when they cannot be certain that they are not carrying and spreading the virus. Our findings could be used as an effective “tool” to demonstrate such a cost, clearly presented in terms of number of excessive infections. During these days of uncertainly, we believe it is fundamental to provide scientific facts that could illuminate this connection and make it tangible for the general. The amplitude of the cycles we describe here is such that many thousands of infections could be averted by carefully scrutinizing local policies, medical practice, and social norms.

## Introduction

On May 25^th^, 2020, there were over 5.5 million reported cases of SARS-COVID-2 (COVID-19) worldwide, and over 346,000 deaths^1^. In many countries social distancing guidelines and/or restrictions have been put in place to limit the spread of the disease. Yet, there seems to be a world-wide tendency to negate some of these restrictions, at least within certain subpopulations. In a few countries no formal restrictions have been imposed on social interaction. Importantly, the pool of people with whom social interactions occur may depend on the specific day of the week. During working days, most interactions may occur within a stable pool of people, namely household members and/or co-workers, whereas during weekend days, a different and more diverse pool of people may apply, including parents, other relatives, and the extended social network.

The price of not imposing restrictions on social interactions, or partially or completely defying them, is ambiguous, although it is assumed that those who do not maintain social distancing are at higher risk of getting infected and spreading COVID-19. Also it is unknown whether temporary or partially loosening of restrictions, specifically during a weekend or a holiday (by the entire population or a subset of it), or of extending the pool of people with whom we interact during the weekend, has a significant impact on the number of infected people or deaths.

To address these questions, we scanned Our World in Data database^1^ (derived from the European Center for Disease Prevention and Control (ECDC) data), as well as specific databases of states within the USA, for (i) daily new cases, (ii) daily numbers of hospitalizations (when available), and (iii) daily deaths. We observed marked weekly rhythmicity in all three indices. To test if the observed patterns are not random, and whether they follow a 7-day cyclic rhythm, we studied all 12 developed North American and European countries that reported more than 1,000 deaths by May 4th. Because some databases ascribe events to the day the information becomes available, rather than to the actual date of event, we also collected data from resources that strictly ascribe events to their occurrence date. To this end we analyzed such authenticated data from 13 states within the USA that had more than 1000 death and clearly indicated that the data is authenticated to the date of event occurrence (of whom 11 provided data on new cases, 2 on hospitalization, and 11 on deaths).

## Results

We analyzed data from March 29 to May 4^th^ (±3 days to create a 7-day moving average). Non-authenticated databases used were Worldometer and ECDC, assessing all 12 European and North-American countries having more than 1,000 deaths by May 4^th^. Authenticated databases were all those we were able to locate in US states that ascertained date-authenticated data and had more than 1,000 deaths by May 4^th^. Table 1 summaries all results for both date-authenticated and non-date-authenticated databases.

**Table 1:** A summary of all data collected.

| Authenticated<br>(Yes/No) | Location | No. of<br>Cases | Sig. 7-day<br>Patter<br>n -<br>Cases?<br>(Yes/<br>No) | Peak<br>Days<sup>1</sup> | No. of Other<br>events | Sig.<br>7-Day<br>Pattern?<br>(Yes/No<br>) | Peak<br>Days | Data-<br>base |
| --- | --- | --- | --- | --- | --- | --- | --- | --- |
| Yes (only<br>Deaths) | Florida | NA | NA | NA | Deaths: 1,343 | No<br>(p=0.069<br>) | Tu, Th-F | State<br>Database |
| Yes | Maryland | NA | NA | NA | Deaths: 1,332 | No | M, W, F | State<br>Database |
| Yes | NYC | 129,117 | Yes***<br>* | M-Th | Deaths: 13,736 | No | NA | City<br>Database |
|  |  |  |  |  | Hospitalization:<br>34,467 | Yes**** | M, Tu |  |
| Yes | Ohio | 99,732 | Yes***<br>* | M-Tu,<br>F | Deaths: 1,273 | No | NA | State<br>Database |
|  | Ohio [0-69] | 7,108 | Yes***<br>* | M, W,<br>F | NA |  |  |  |
|  | Ohio [70+] | 2,853 | Yes***<br>* | M-Tu |  |  |  |  |
| Yes | New Jersey | 98,400 | Yes***<br>* | M-W | NA | NA | NA | State<br>Database |
| Yes | Massachusetts | 64,829 | Yes***<br>* | Th-Sat | Deaths: 4,444 | No<br>(p=0.069<br>6) | M, W-Th | State<br>Database |
|  |  |  |  |  | Average percent<br>of positive tests:<br>23.5% | No | NA |  |
| Yes | Michigan | 32,289 | Yes***<br>* | M-W, F | NA |  |  | State<br>Database |
| Yes | Connecticut | 27,496 | Yes***<br>* | M-Th | Deaths: 2,600 | No | NA | State<br>Database |
| Yes | Georgia | 25,067 | Yes***<br>* | M-Tu,<br>Th | Deaths: 1,274 | No | NA | State<br>Database |

| Yes | Virginia | 21,294 | Yes***<br>* | M, F | Deaths: 741 | No | NA | State Database |
| --- | --- | --- | --- | --- | --- | --- | --- | --- |
|  |  |  |  |  | Hospitalization: 2,506 | Yes**** | M |  |
| Yes | Indiana | 19,456 | No | NA | Deaths: 1,214 | Yes* | Tu | State Database |
| Yes | Colorado | 14,206 | Yes***<br>* | M, F | Deaths: 1,024 | No | NA | State Database |
| Yes | Washington | 9,541 | Yes***<br>* | M, Th-F | Deaths: 480 | No | NA | State Database |
| Authenticated (Yes/No) | Location | No. of Cases | Pattern significant ? (Yes/No) | Peak Days<sup>1</sup> | No. of Other Events | Pattern significant? (Yes/No) | Peak Days | Data-base |
| No | US (All states) | 1,088,047 | Yes**** | Th-Sat | Deaths: 67,167 | Yes**** | Tu-W | Worldometer |
| No | United Kingdoms | 172,056 | No | NA | Deaths: 27,285 | Yes**** | W, F-Sat | Our-World-in-Data |
| No | Spain | 139,060 | Yes**** | W-Sat | Deaths: 20,570 | No (p=0.09) | W-F | Our-World-in-Data |
| No | Italy | 124,219 | Yes*** | Th-Sun | Deaths: 19,748 | Yes*** | W, F, Sun | Our-World-in-Data |
| No | Germany | 114,593 | Yes** | Th-Sat | Deaths: 6367 | Yes*** | W-F | Our-World-in-Data |
| No | Canada | 54,799 | No | NA | Deaths: 3,629 | Yes**** | F-Sun | Our-World-in-Data |
| No | Belgium | 42,622 | No | NA | Deaths: 7,555 | No (p=0.067) | NA | Our-World-in-Data |
| No | Netherlands | 31,968 | Yes**** | F-Sat, M | Deaths: 4,510 | Yes** | W-F, Sun | Our-World-in-Data |
| No | Sweden | 19,271 | Yes* | Th-Sat | Deaths: 2,587 | Yes**** | W-F | Our-World-in-Data |
| No | Switzerland | 17,718 | Yes* | F, Sun | Deaths: 1,275 | Yes* | W-Th, Sat-Sun | Our-World-in-Data |
| No | Austria | 7,841 | Yes * | W-Th, Sat | Deaths: 530 | No | NA | Our-World-in-Data |
| No | Denmark | 7,477 | No | NA | Deaths: 432 | Yes* | Sun | Our-World-in-Data |

All databases above were accessed between May 20<sup>th</sup> to May 22<sup>nd</sup>, and all data (new cases, deaths, hospitalizations, and %of positive tests) were obtained for March 29<sup>th</sup> to May 4<sup>th</sup> 2020.
<sup>1</sup> Peak days were selected based on the days that showed most new cases numbers each week (compared to a seven-day running average). Only days that ranked in the three first places three times or more, of the five weeks, were selected. Days are presented in abbreviated form (M=Monday, Tu=Tuesday, W= Wednesday, Th=Thursday, F=Friday, Sat=Saturday, and Sun=Sunday)
\* - p-Value is less than 0.05
\*\* - p-Value of less than 0.005
\*\*\* - p-values is less than 0.0005
\*\*\*\* - p-Value of less than 0.00005

### New cases and hospitalization

We first analyzed non-authenticated data from Worldometer regarding US as a whole, and found a significant 7-day periodicity in new cases, having a sinus shape, peaking on Fridays and decreasing toward a nadir on Monday (Figure 1A). Within the additional 11 European/North-American countries, very similar patterns were observed in 8 of them (Table 1, and Supp. Fig 1 for representative countries that show this periodicity).

**Fig. 1:**
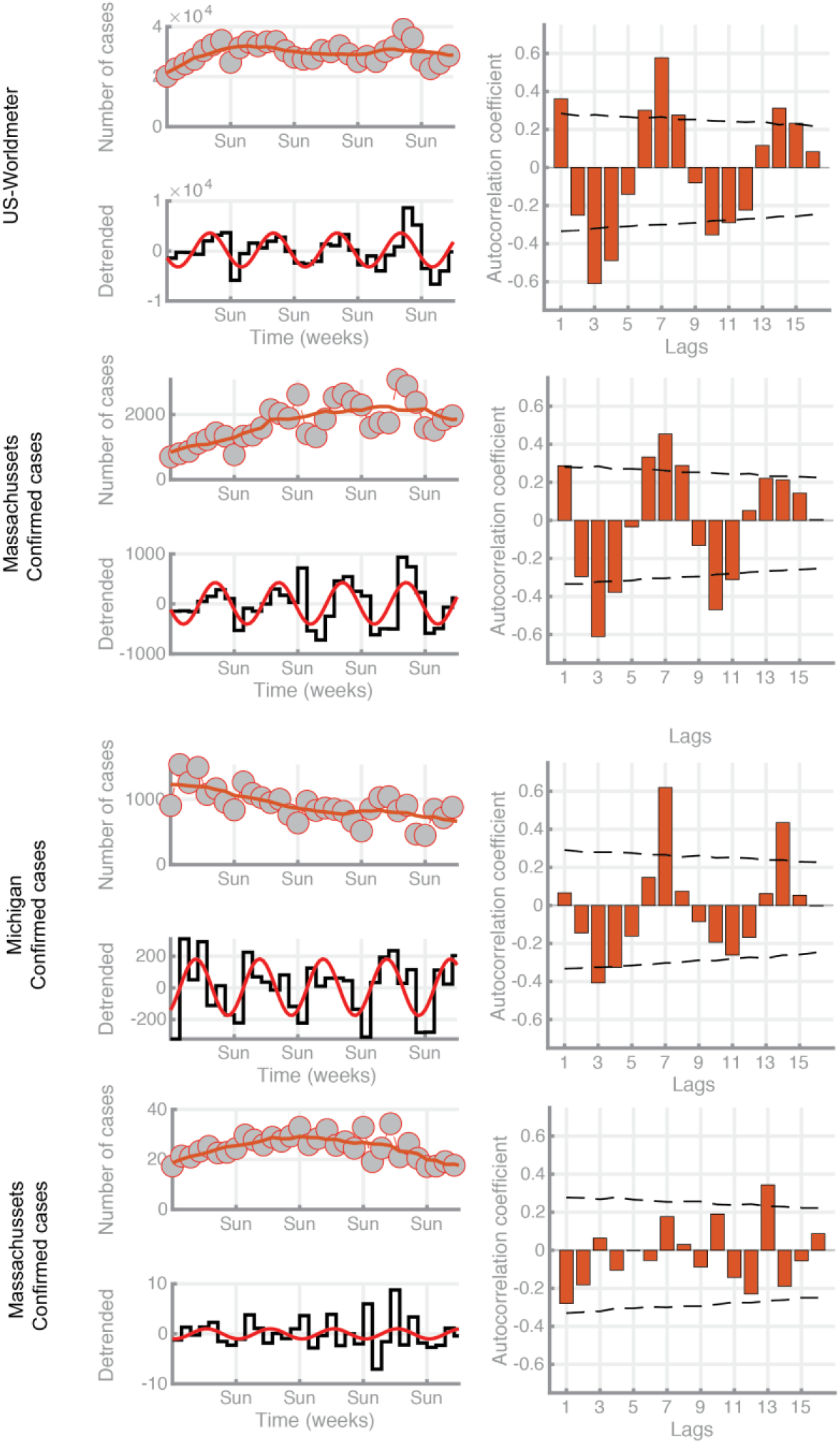
US data representative outcomes of the entire US (not date-authenticated, based on the Worldometer database) and two representative date-authenticated states (Massachusetts and Michigan) The upper left pane depicts raw data (gray points) with averaged data over-imposed (red line). The lower left pane presents the detrended data (difference between the raw data and a 7-day moving average - 3 days before to 3 da s after each data point, black line) with a fixed 7-day periodic sinusoidal superimposed (in red). The right pane presents autocorrelations for lags ranging between 1 and 16 days, with 95% confidence intervals (calculated based on bootstrap resampling analysis). Significant autocorrelations fall outside the 95% interval

We then analyzed data from US states that clearly indicated that the information provided is date-authenticated. Eleven states provided such data on new cases (see a complete list in Table 1), and in 10 of them a significant periodic seven day cycle was observed (See Fig. 1-3 and Supp. Fig. 2-4 for representative states data), commonly peaking on weekdays, 2-6 days after the weekend. In Massachusetts, the databases also included the percent of positive tests/day, which did not show periodicity and ranged between 10%-30% along the entire period studied.

**Fig. 2:**
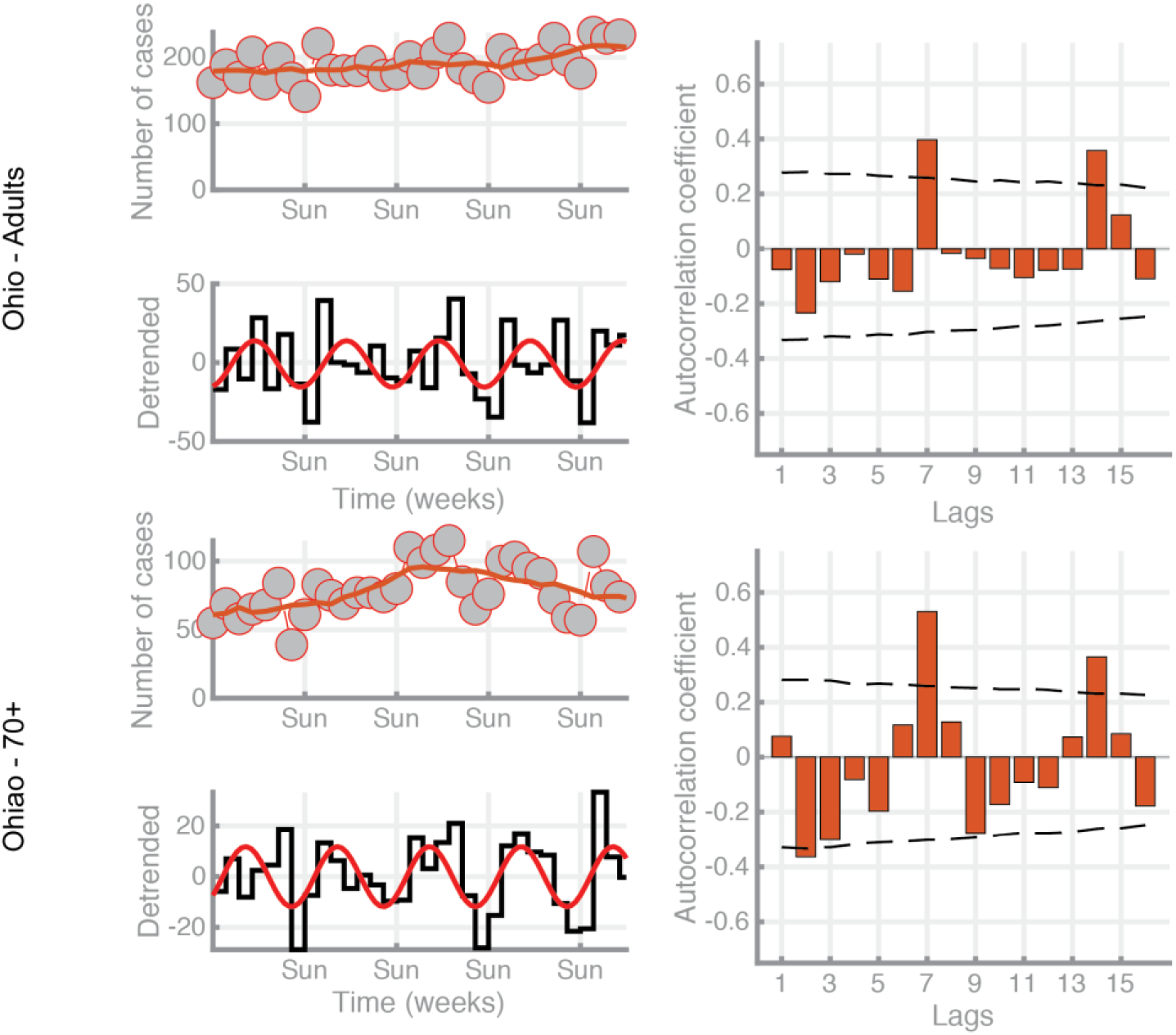
Ohio New Cases by age graphs: A. Age below 70, B. Age over 70 (Date-authenticated. Date is defined as illness onset or if onset date in unknown, the earliest known date associated with the case). For explanation of depicted data, see Fig. 1 legend.

In New York City we located date-authenticated data for new cases, hospitalization, and deaths. Both new cases and hospitalizations exhibited a significant 7-day periodicity, while deaths showed a statistically non-significant pattern for a 7-day periodicity (Fig 3). Hospitalizations also showed a 7-day periodicity in Virginia, summing up to 2 of 2 states where this authenticated information was available.

**Fig. 3:**
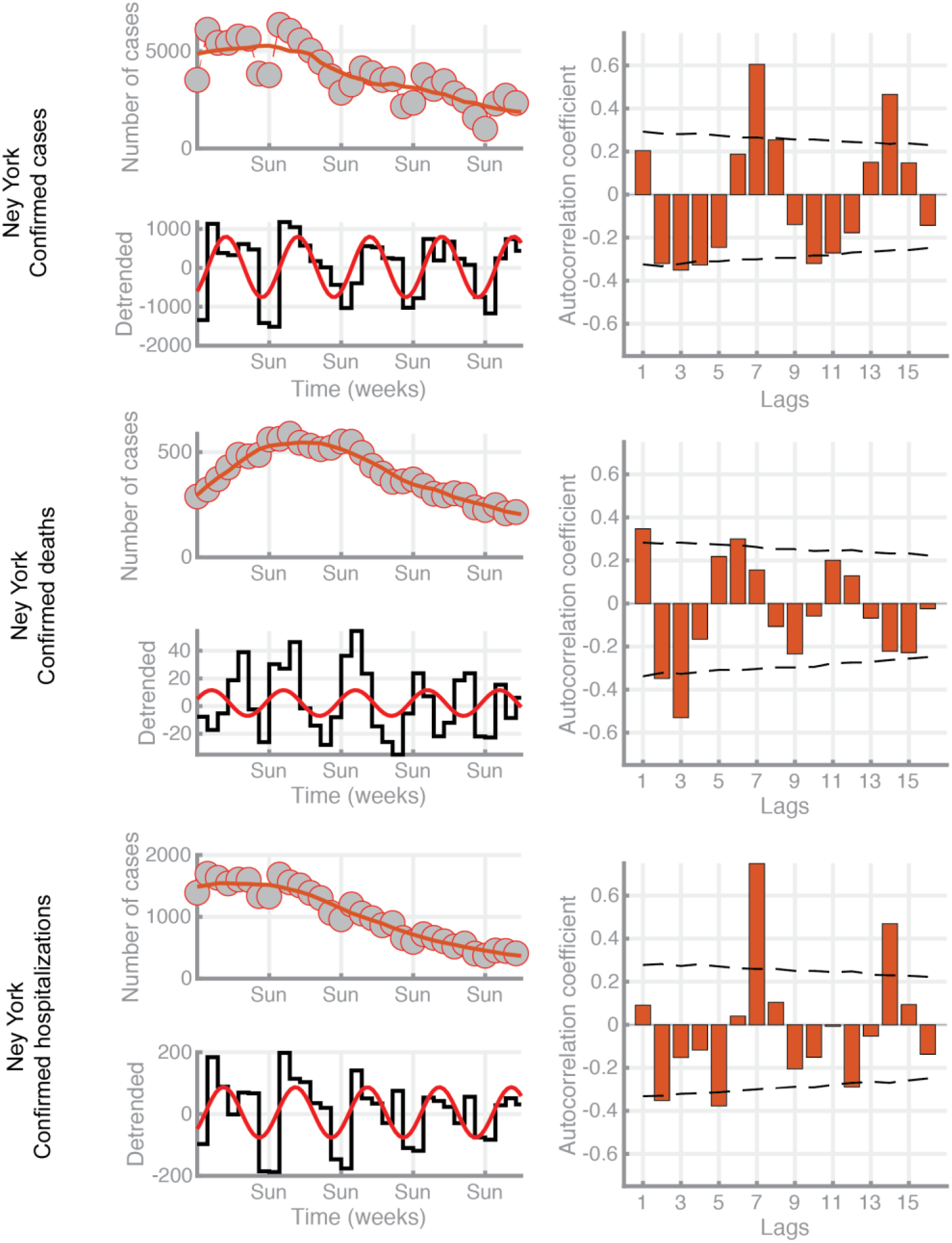
NYC: New Cases, Hospitalization, and Death Data (Date-authenticated). For explanation of depicted data, see Fig. 1 legend.

**Fig. 4:**
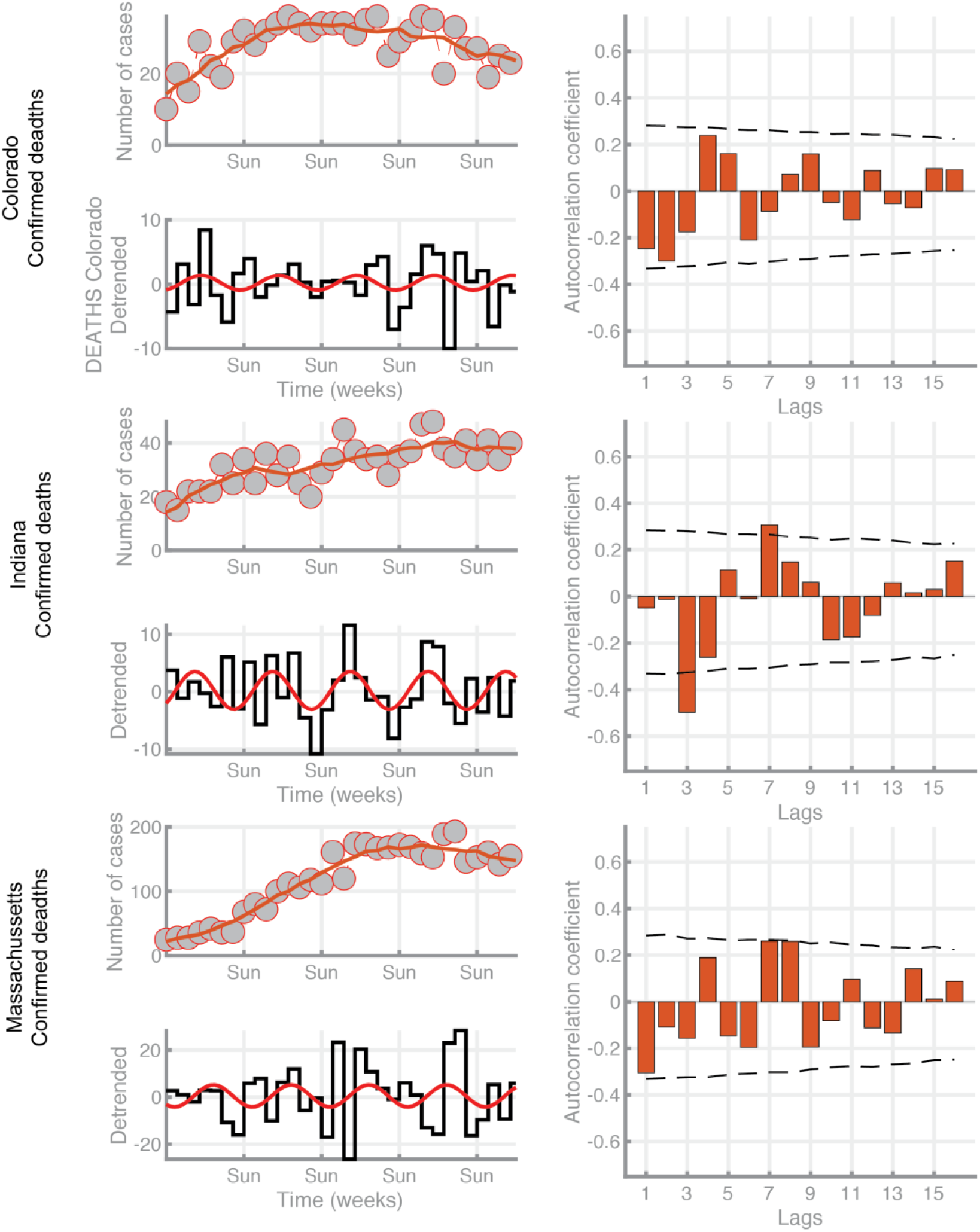
COVID-19 Deaths in the US: representative date-authenticated states : Indiana, Colorado, and Massachusetts. For explanation of depicted data, see Fig. 1 legend.

### Death patterns

Based on non-authenticated databases, of the 12 European/North-American countries studied, 9 presented a significant 7-day periodicity (See Fig. 1 and Supp Fig 1 for examples). Within the 11 US states providing date-authenticated data on death date, only 1 presented a statistically significant 7-day periodicity, 2 additional states presented a noticeable but non-statistically-significant 7-day periodicity, and 8 did not present any consistent periodicity.

## Discussion

Worldmeter database showed a significant 7-day periodicity in the US, both regarding new cases that peak on Fridays, and deaths that peak on Tuesdays/Wednesdays. Based on ECDC non-authenticated data for North-American/European countries, a 7-day cyclic pattern was evident in 8 out of the 12 countries studied, regarding new cases, and in 9 out of the 12 countries regarding number of deaths, with peak incidence of new cases occurring mostly on Thursdays-Saturdays, and peak incidence of death occurring mostly on Wednesdays-Fridays.

More important is data from date-authenticated US states that showed statistically significant 7-day periodicity patterns. Specifically, a significant 7-day pattern was found in 10 of 11 states regarding new cases, 2 of 2 regarding hospitalizations, and 1 of 11 regarding deaths.

Several characteristics of the COVID-19 pandemic and disease course are known and potentially hold relevance to these observations and to our speculated interpretations. First, the disease is commonly transmitted through respiratory droplets during un-protected social interactions^3^. Second, older people account for most recognized cases of the disease, and for the great majority of fatalities^4^, and asymptomatic people carry and transfer the virus^5^. Third, epidemiological studies indicated that the median duration between contacting a sick/carrying individual and exhibiting clinical symptoms of COVID - 19 infection is 4-5 days (50% up to 5 days)^5^. Also relevant to our hypothesis, a trend towards increased social mixing during the weekends has been reported (including in countries present in our analyses^6^), and weekend interactions are expected to involve a different pool of people than weekday work & households interactions.

Because the restrictions imposed in many countries or adopted voluntarily include limiting or eliminating workplace hours/interactions, at least for people at high-risk, we hypothesize that susceptible/older people and retired people may become infected at higher rates during weekend-days compared to weekdays, as a result of increased social interactions with younger relatives or friends during the weekend^6^, or as a result of interacting with a different and extended pool of people. Under this assumption, and in accordance with the above known characteristics of COVID-19, it follows that a significant portion of these vulnerable individuals will exhibit clinical signs of COVID-19 infection at higher rates ∼4-5 days after the weekend, on Wednesday-Friday. The pattern regarding deaths is expected to exhibit greater variance due to the prolonged lag from infection to death (2-3 weeks from symptoms onset to death), and a variety of medical and environmental intervening variables. Both predictions are indeed evident in our results of date-authenticated data. A higher infection rate during weekend days may stem from lower social distancing, or higher frequency of interactions between young and old individuals during the weekend. Furthermore, as most new symptomatic cases of COVID -19 occur in people older than 70, and the majority of deaths are within this age group, we believe that this vulnerable population is infected by unaware younger relatives or friends carrying the virus. It is also likely, but need to be verified, that this vulnerable population exhibits shorter and more consistent lags of time from infection to symptom onset and to death.

Alternative explanations for the seven-day cycle in the numbers of reported new cases and reported COVID-19 mortality should be considered. A leading hypothesis would be a selective delay in reporting of these events. Specifically, one may suggest that while new cases and deaths are distributed evenly along all 7 days of the week, reports of these events are delayed during the weekend, and erroneously ascribed to 2-4 days later, whereas events occurring on weekdays are reported without delay. In the USA, however, we were able to locate 11 states with more than 1000 deaths that indicate the actual date of new cases, as defined by the actual day of symptoms onset, or the date a positive test was taken (see Table 1). Hospitalization (2 states) and death rates (11 states) were also logged accurately and authenticated. In 10 of these 11 states, the seven-day cyclic pattern is clearly evident regarding new cases, and in 2 of 2 states the phenomenon is observed regarding hospitalization, refuting a delayed report as an alternative explanation for a weekly cycle in new cases. One may nevertheless argue that people that present symptoms of COVID-19 on the weekend may delay taking a test (or reporting symptom onset) to Monday/Tuesday, in contrast to those that present symptoms on weekdays, who would not delay it. This scenario will yield a marked peak of new cases on Monday/Tuesday, decreasing towards Friday, which is not evident. We also checked for the percent of positive tests along the entire week in Massachusetts (where this information was available and a 7-day cycle was significant), and found no significant cycles in this index, nor a shortage in tests during weekends.

Weekly cycle pattern in death may stem from previous periodicity in infection dates, and is unlikely to be attributed to individual choice, as most patients are anesthetized long before dying. Of the 11 date-authenticated sates in the US, this pattern was significant only in one (Indiana), an evident but not statistically significant in additional 2 states (Massachusetts and Florida). The effect sizes for death seem smaller than for new cases and hospitalization. This less profound cycle would be expected given the longer delay from infection to death (average estimation of 2-3 weeks), the expected greater variance in developing severe symptoms leading to death, and the smaller number of events. Thus, the more pronounced weekly cycles in 9 of the 12 countries based on ECDC data may indeed be attributed, at least to some extent, to weekends delay in reporting deaths. However, the smaller but statistically significant effects in the date-authenticated states in the USA suggest that this cycle is a remnant consequence of weekend high-infection rates.

If our above postulations indeed account for the observed 7-day patterns, many lives may be saved by greater adherence to social/physical distancing, especially during the weekend. Alternatively, such adherence may at least flatten the curves of hospitalization and mortality, which in some countries with restricted medical resources would also lead to saving lives. In the 11 states in the US, where date-authenticated data was used, the cumulative low range of daily new cases in the period assessed is about 10,000, while the high range is about 15,000. If the peak days are truncated, a weekly saving of ∼20,000 new cases in the 11 states studied is expected. Notably, the potential of truncating weekly peaks should be weighed against the expected mental and medical costs of loneliness and isolation as a consequent of greater adherence to social isolation.

Our hypothesis should be re-tested and refined based on more complete worldwide databases that accurately report events’ times, if and when these will become available. We hope that our analysis will trigger local health authorities to ascertain or improve the accuracy of the reported dates, and act accordingly to prevent unnecessary spread of COVID19 during weekends.

## Methods

### Statistics

Analyses were performed separately for each country/US state for daily numbers of new cases, hospitalizations, and for deaths. In order to account for global trends in each country/state, a seven-day moving average (from 3 days before to 3 days after) was calculated and the detrended data (i.e. the residuals between actual data and moving average) used for further analyses. Next, autocorrelations were calculated (implemented in SPSS, Version 25), using lags ranging between 1 and 16 days. An autocorrelation is the correlation of a signal with a delayed copy of itself as a function of a given lag. The results represent the similarity between observations as a function of the time lag between them. As such, a significant autocorrelation indicates the presence of a recurring pattern such that values in the series can be predicted based on its preceding values. For example, if an autocorrelation with a lag of 7 days is positive and significant, it means that the values of the series tend to repeat themselves every seven days. We hypothesized that autocorrelations with a lag of 7 days will be significantly positive. Significance and confidence intervals were calculated based on a simulated Bootstrap resampling analysis. For this purpose, within each dataset, data-points were randomly selected with replacement from the original pool and autocorrelations were calculated 10,000 times.

Confidence intervals were obtained by using the 95% central values of the simulated autocorrelations. P-values were calculated by counting the number of simulated autocorrelations that were more extreme than the original autocorrelations, and dividing by 10,000. Finally, for the purpose of comparison of the detrended time series with a fixed 7-day periodic sinusoidal pattern, we fitted the following model *y* (*t*) *= b*_1_ * (sin (2π*t*/*c* + 2π*t*/*c* + *b*_2_)) + *b*_3_ where b1 represents the amplitude, b2 phase shift and b3 an offset while c was held constant to match the desired period of 7 days. The fit was computed with a simplex search method as part of the Matlab Optimization Toolbox (Matworks Inc).

## Data Availability

All data is availble through Our World In Data and Worldometer websites.
Data regarding specific US states is available through links showed in Table 1.

https://ourworldindata.org/coronavirus

https://www.worldometers.info/coronavirus/#countries

## Author Contirbutions

**Itay Ricon-Becker and Shamgar Ben-Eliyahu**: literature search, figures, study design, data collection, data interpretation, writing. **Ricardo Tarrasch and Pablo Blinder**: literature search, figures, study design, data collection, data analysis, data interpretation, and writing.

## Competing interests

None

## Funding

None

## Figures

**Supp Fig 1:**
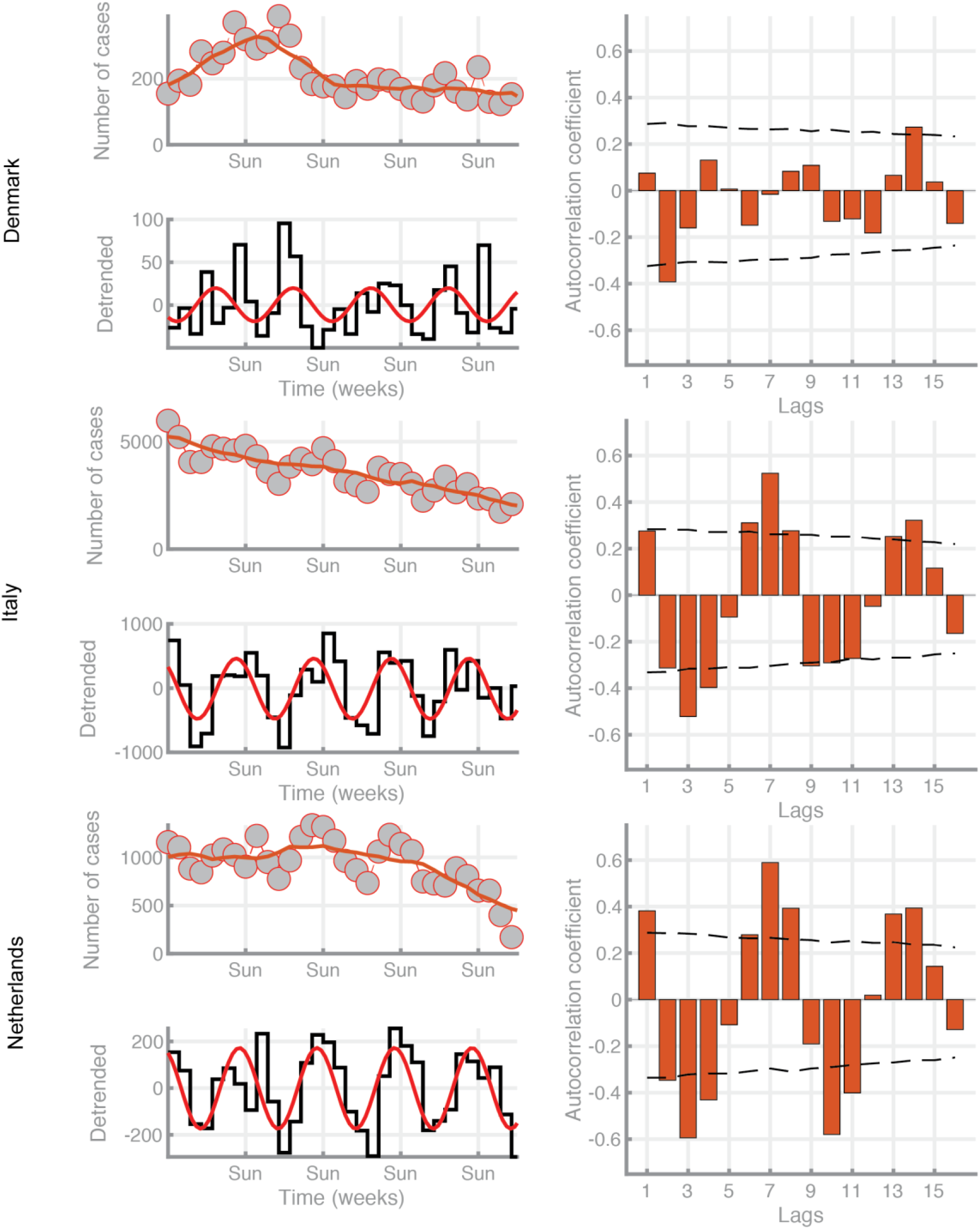
New Cases: Representative not date-authenticated countries data: Italy, Netherlands and Denmark. For explanation of depicted data, see Fig. 1 legend.

**Supp Fig 2:**
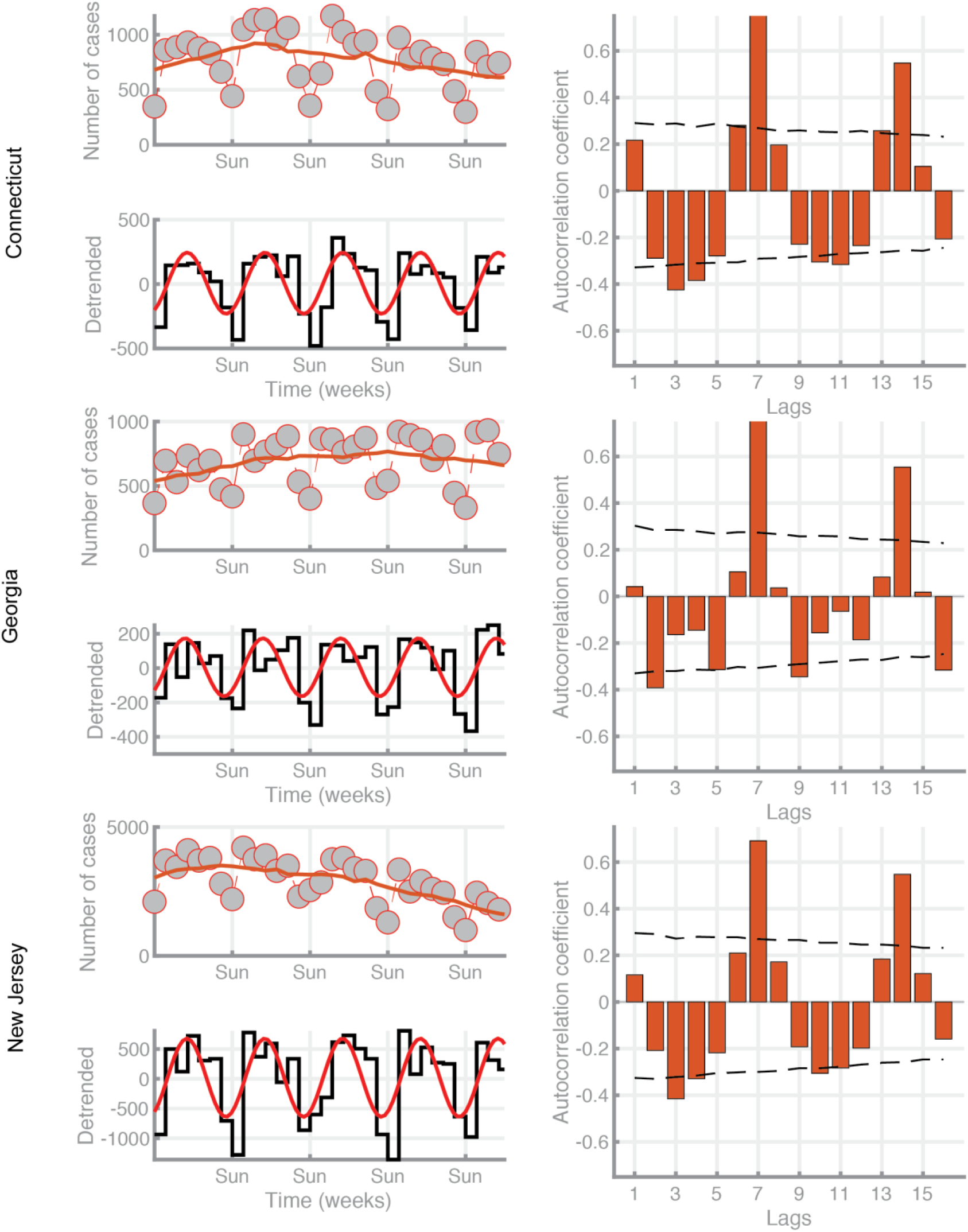
New Cases in representative date-authenticated states data: Connecticut. Georgia, New Jersey. For explanation of depicted data, see Fig. 1 legend.

**Supp Fig 3:**
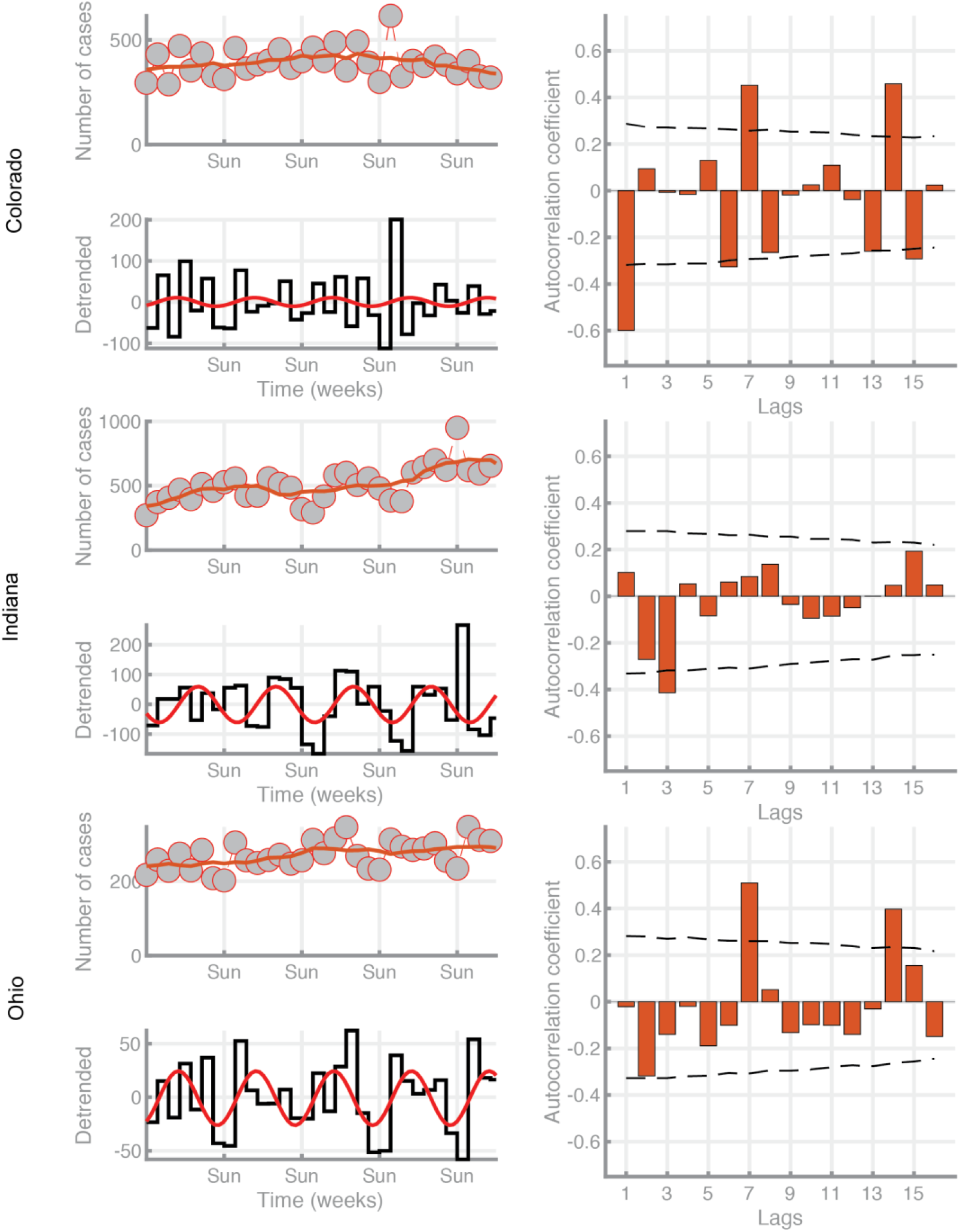
New Cases in representative date-authenticated states data: Ohio, Colorado, Indiana. For explanation of depicted data, see Fig. 1 legend.

**Supp Fig 4:**
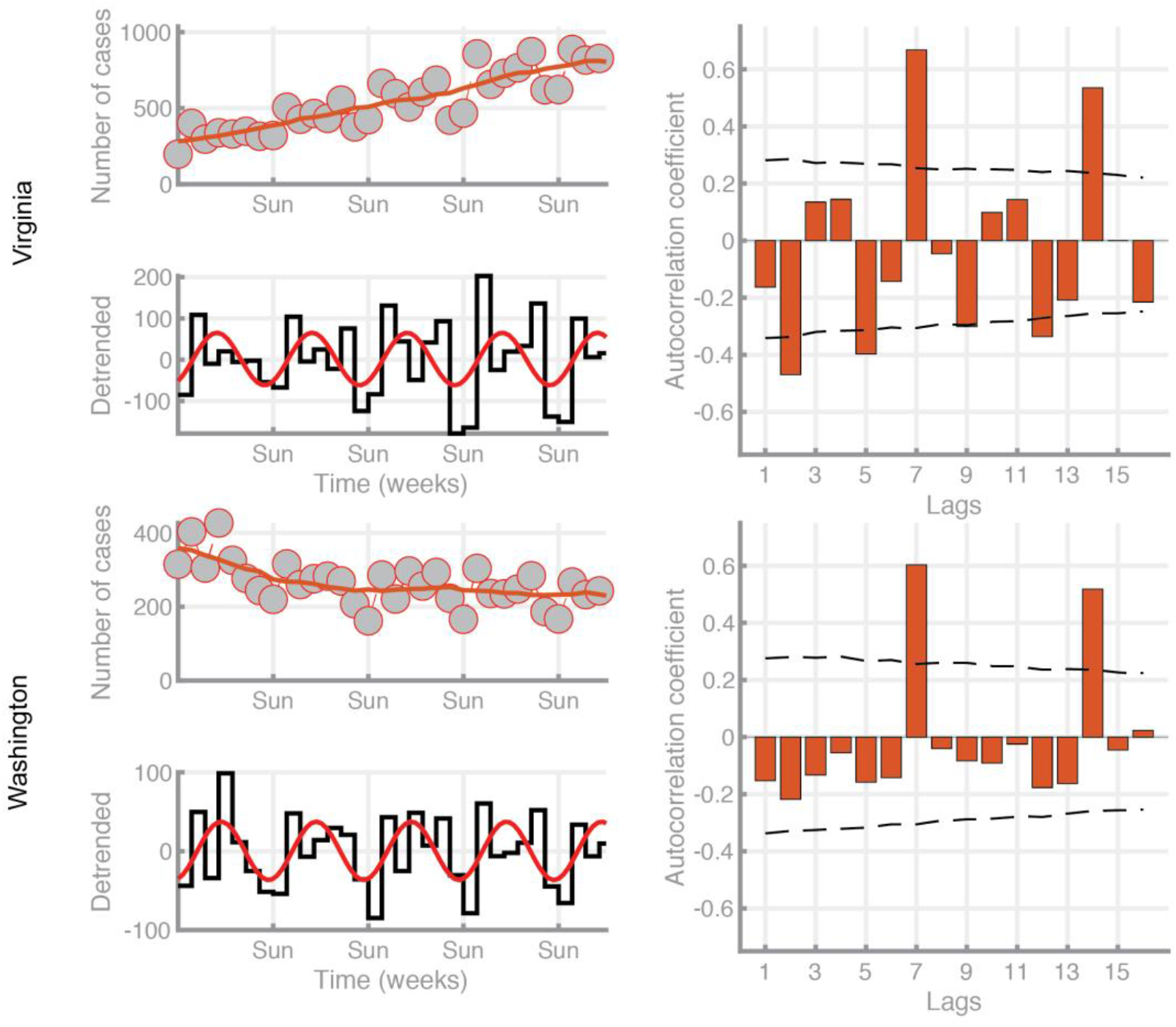
New Cases in representative date-authenticated states data: Virginia, Washington. For explanation of depicted data, see Fig. 1 legend.

## Notes

### Competing Interest Statement

The authors have declared no competing interest.

### Funding Statement

No external funding was received

### Summary of Updates

The paper was revised in order to add a more in-depth analysis of data from specific US states that provide data with greater accuracy of reporting. Table 1 was added, and the graphs were also edited in order to introduce these new analyses.

